# Effects of COVID-19 lockdown on heart rate variability

**DOI:** 10.1101/2020.07.30.20165118

**Authors:** Nicolas Bourdillon, Sasan Yazdani, Laurent Schmitt, Grégoire P.Millet

## Abstract

**Introduction:** Strict lockdown rules were imposed to the French population from 17 March to 11 May 2020, which may result in limited possibilities of physical activity, modified psychological and health states. This report is focused on HRV parameters kinetics before, during and after this lockdown period.

**Methods:** 95 participants were included in this study, who underwent regular orthostatic tests (a 5-minute supine followed by a 5-minute standing recording of heart rate (HR)) on a regular basis before, during and after the lockdown (BSL, CFN and RCV, respectively). HR, power in low- and high-frequency bands (LF, HF, respectively) and root mean square of the successive differences (RMSSD) were computed for each orthostatic test, and for each positions. Subjective well-being was assessed on a 0-10 visual analogic scale (VAS).

**Results:** Out of the 95 participants, 19 (WB+) reported an improved well-being (i.e., increase >2 in VAS score) during CFN, contradictory to the 76 other participants (WB-). There was an increase in HR and a decrease in RMSSD when measured supine in CFN and RCV, compared to BSL in WB-, whilst opposite results were found in WB+ (i.e. decrease in HR and increase in RMSSD in CFN and RCV; increase in LF and HF in RCV). There was a moderate significant correlation between VAS and HR, RMSSD, HF, respectively, in the supine position; the higher the VAS score (i.e., subjective well-being), the higher the RMSSD and HF and the lower the HR. In standing position, HRV parameters were not modified during CFN.

**Conclusion:** The strict COVID-19 lockdown likely had opposite effects on French population as 20% of participants improved parasympathetic activation (RMSSD, HF) and rated positively this period, whilst 80% showed altered responses and deteriorated well-being. The changes in HRV parameters during and after the lockdown period were in line with subjective well-being responses. These results confirmed the usefulness of HRV as a non-invasive means for monitoring well-being and health in the general population.

## INTRODUCTION

The COVID-19 pandemic in 2020 resulted in strict confinement rules in France for 8 weeks, from 17 March to 11 May, period known as the lockdown. Its aim was to slow down the spread rate of COVID-19, therefore saving lives and decreasing the workload in hospitals, which capacity to treat new patients was exceeded. Restrictions during the lockdown included closed gyms and sports centers, plus limitations on the duration spent outdoors, maximum walking distance from home which, associated with the lack of space and appropriate devices in homes for physical exercise, and lack of technical knowledge of the population on appropriate training routines, resulted in decreased physical activity [1]. A survey by the French public health national institute “Santé Publique France” published on 17^th^ June 2020 confirmed that 47.4% of the population declared a decrease in physical activity, especially (58.9%) in walking time, during the lockdown period. Contradictory, a minority (17.9%) used this period for increasing their physical activity. In addition, 33.4% of the population declared spending more than 7h seated. The average daily seated time was 6h19. Beyond these self-declared data, the impact of the lockdown period remains unclear, but one may hypothesize that it may have led to changes in cardiovascular fitness and psychological states [2].

Exercise plays a fundamental role in cardiovascular health [3] but also in mitigating the psychological impacts of lockdown [4]. Heart rate variability (HRV) is a commonly used method for cardiovascular follow-up in athletes [5]. Specifically, HRV analysis allows to evaluate the modulation of the sympathetic and parasympathetic branches of the autonomic nervous system. The low-frequency (LF) band reflects a mix of sympathetic and parasympathetic modulations on the heart [7] with likely a sympathetic preponderance. The migration of the respiratory sinus arrhythmia in the low-frequency band is a subject of debate [6]. The sympathetic modulations seen in LF are likely due to the vasomotor tone or to the central modulation of the sympathetic tone [5], which is linked to changes in arterial blood pressure [8] and baroreflex activity [9]. Respiratory sinus arrhythmia is the main phenomenon provoking changes in the HF band which therefore mainly reflects parasympathetic influences on the heart [10]. Physical exercise increases the parasympathetic tone [11]. The best performance occur with hypertonia of both the sympathetic and the parasympathetic nervous systems [12,13]. Hypotonia of one or the other is a sign of cardiovascular deconditioning [14]. Finally, HRV-based methods have proven valid and reliable in the evaluation of stress, well-being and recovery [15].

To the best of our knowledge, HRV parameters before, during and after the lockdown period have not been reported in the literature. Therefore, the aim of the present study was to report HRV parameters to demonstrate the effects of lockdown and to investigate whether changes in HRV were related to changes in psychological states during and after this period. We hypothesized that alteration in parasympathetic-related markers (i.e., decreases in RMSSD and HF, increase resting HR) would reflect a negative feeling over the period, whilst a minority of the population with a positive appreciation of this particular period would report an improved HRV.

## METHODS

### Experimental design

Strict lockdown (further denoted CFN) rules applied in France over 8 weeks, from 17 March to 11 May 2020. The lockdown was unexpected, therefore there was no previous intention to study its effects on HRV. We used HRV monitoring setup dedicated to other studies to collect data. Data collection was therefore performed for the purpose of this study. Three periods were compared: From 1 January to 16 March (baseline, BSL); from 17 March to 11 May (CFN), and from 12 to 31 May (recovery, RCV). The local ethical committee approved the study (agreement 2016-00308; Commission Cantonale d’Ethique de la Recherche sur l’être humain, CCER-VD; Lausanne, Switzerland) as part of a set of studies on a broader scale. All experimental procedures conformed to the standards set by the Declaration of Helsinki

### Participants

The participants in this study were equipped with HRV monitoring devices before the beginning of lockdown. Inclusion criteria were, age between 18 and 60 years old, physically active, healthy, non-smokers, with no known diseases and no pregnancy or lactation for women, living in France. Out of the database (n = 112), participants with at least forty orthostatic tests between 1 January and 31 May 2020 and at least 10 tests in each period of three periods (BSL, CFN and RCV) were extracted (n = 95).

### Heart rate variability

The participants performed an orthostatic test (5 minutes supine followed by 5 minutes in the standing position) on a regular basis between the 1^st^ of January 2020 and the 31^st^ of May 2020 [16]. This orthostatic test had to be performed, in the morning, upon wake-up, with an empty bladder, and before breakfast. The inter-beat interval (RR interval) measuring device (sensor H7 + chest belt, Polar, Kempele, Finland or cardiosport TP5+, Cardiosport, Waterlooville, UK) was connected to the participants’ smartphones via Bluetooth (mobile application: inCORPUS^®^ v2.1.3, be.care SA, Renens, Switzerland). Data were immediately and automatically transferred to our servers for analysis. Therefore, meetings between participants and researchers were avoided, in the respect of the strict lockdown rules applying.

The RR intervals from the orthostatic tests were first analyzed to remove ectopic beats from the recordings. Ectopic beats were then compensated for by means of interpolation to calculate normal-to-normal (NN) intervals. From the NN intervals, the following heart rate variability (HRV) parameters were extracted: mean HR; the root mean square of the successive differences (RMSSD); the spectral power in the low-frequency (LF, 0.04 – 0.15 Hz) and high-frequency bands (HF, 0.15 – 0.40 Hz) in ms^2^; and the values (expressed in normalized units) for LF and HF, labelled nLF and nHF, respectively [5,17]. The spectral power was estimated using a fast Fourier transform on the resampled NN intervals (4 Hz) using a window length of 250 data points and an overlap of 50%. All computations were performed separately for the supine (SU) and standing (ST) positions using. All analyses were performed using MATLAB^®^ (R2019a, MathWorks, Natick, MA, USA).

### Visual analogic scale (VAS)

On the mobile application, for each test, the participants were required to rate their well-being “how do you feel today?” using a visual analogic scale (VAS) graduated from 0 to 10.

### Statistics

All results are given as mean ± SD. The Kolmogorov-Smirnov test was used to ensure normality of the data. Measurements were evaluated with a two-way ANOVA analysis for time effect (BSL, CFN and RCV) and group effect (WB- vs. WB+). The statistical power of the performed tests was set at p = 0.05 for significance and p = 0.10 for tendency. The Tukey-Kramer post-hoc was used when appropriate. Pearson’s coefficient correlations and significance were computed for LF, HF, HR and RMSSD against VAS, in the supine and the standing positions. Statistical analyses were performed using MATLAB^®^.

## RESULTS

Ninety-five participants (27 women, 68 men) were included in this report (age 37 ± 11 years, height 176 ± 8 cm, weight 71 ± 12 kg). Out of these participants, 20% (WB+, n = 19; 5 women, 14 men; age 36 ± 13 years, height 178 ± 10 cm, weight 74 ± 13 kg) showed an improved well-being, estimated by the positive change (>2) in the 0-10 VAS score between BSL and CFN. Contradictory, most of the participants (WB-, n = 76; 22 women, 54 men; age 38 ± 11 years, height 175 ± 7 cm, weight 69 ± 12 kg) did not improve or deteriorated this VAS score during the CFN period.

Figure 1 shows the negative impact of CFN in the WB- group, with a general alteration in parasympathetic activity in the supine position, as shown by the increased supine HR and decreased RMSSD in CFN and RCV compared to BSL. There was a tendency for a decrease in RMSSD between CFN and RCV. There was also a decrease in HF in RCV compared with BSL. In the standing position, there was a significant increase in HR in CFN and RCV compared to BSL, but there was no change on the other parameters.

**Figure 1:**
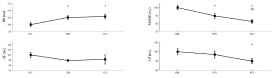
HR, RMSSD, LF and HF before (BSL), during (CFN) and after (RCV) the strict lockdown period in France in 76 participants who deteriorated or maintained their well-being over the CFN period. * p < 0.05 for difference with BSL. # p < 0.05 for difference with CFN.

Figure 2 shows the positive impact of CFN in the WB+ group with a decrease in supine HR in CFN and RCV compared to BSL, an increase in supine RMSSD in CFN and RCV compared to BSL and an increase in RCV compared to CFN. There was also an increase in LF and HF (and therefore total power) in RCV compared to BSL, and an increase in HF in RCV compared to CFN. There was no change in the standing position.

**Figure 2:**
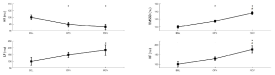
HR, RMSSD, LF and HF before (BSL), during (CFN) and after (RCV) the strict lockdown period in France in 19 participants who improved their well-being over the CFN period. * p < 0.05 for difference with BSL. # p < 0.05 for difference with CFN.

There was an interaction time x group for HR, RMSSD and HF (p < 0.001, p < 0.001 and p <0.01, respectively) in the supine position. There was a significant interaction for HR only (p<0.05) in the standing position. In the supine position, HR was significantly higher in the WB- group compared to the WB+ group in CFN and RCV. RMSSD was significantly lower in the WB- group compared to the WB+ group in CFN and RCV. HF was significantly lower in the WB- group compared to the WB+ group in RCV. In the standing position HR was significantly higher in the WB- group compared to the WB+ group in CFN.

There were significant correlations (p < 0.001 for all) between VAS and HR ((R = -0.24), RMSSD (R = 0.14) and HF (R = 0.19) on the whole population in the supine position. The higher the VAS score (i.e., subjective well-being), the higher the RMSSD and HF and the lower the HR (Figure 3).

When dividing the CFN period in two sub-periods of 4 weeks, there were no differences in any of the parameter between CFN1 and CFN2, suggesting a relative stability across CFN (data not shown).

## DISCUSSION

The present study aimed to investigate the HRV responses of participants exposed to the strict lockdown that occurred in France. We found that HRV changes from BSL to CFN and RCV were contradictory between two groups who did vs. did not deteriorate their subjective well-being over these periods. In 80% of the participants, parasympathetic-related HRV parameters (i.e., RMSSD, HF) measured in the supine position were decreased concomitantly to a psychological alteration. Contradictory, in 20% of the participants, both supine HRV and well-being were improved. Standing HRV was not modified during CFN.

The sudden lockdown that occurred implied drastic changes in the lifestyle of the participants, which is generally a stressor factor. These lifestyles and behaviors included a given level of physical exercise to maintain an adequate health status [18], and guarantee an active aging by reducing the risk associated diseases in older people [19]. Moreover, the psychological impact of lockdown included negative psychological effects, such as frustration, boredom, loss of social contacts, [20] inadequate supplies, inadequate information, financial loss, infection fears, confusion, and anger [21,22]. The participants in the present study lived in France, where a survey of the French national public health institute (Santé Publique France) demonstrated a drastic decrease in physical activity and increased time spent in the seated position [2]. In the present study, physical activity was not measured, but only subjective feeling of well-being, measured by a VAS with a smartphone. This resulted in 20% of the participants who rated positively the lockdown period, since VAS score improved by at least 2 on the 0-10 scale. Interestingly, this is in line with data from the French national public health institute, who reported that 18% of the French population declared an increase in physical activity. Obviously, well-being can be determined by many causes and physical activity is one of them.

In the present study, 80% of the participants reported an increased HR associated to decreased RMSSD and increased LF: these changes could be the consequences of a cardiovascular detraining, with decreased parasympathetic influences on the heart. The loss of outdoor activities as walking is likely of clinical importance since lockdown accelerates vicious circles between isolation, lack of physical activity, psychological stress/anxiety and impaired immunity [23]. Meanwhile, the participants had more time to exercise, but they had to do it by themselves, based on their personal knowledge and material possibilities, with extremely limited external advises and totally absent group motivation. According to the French national public health institute, only 18% of the French population was able to increase the physical activity during the lockdown period. This likely resulted in only 20% of the participants in the present study able to maintain or improve their HRV parameters as shown on figure 2. In this group, the fact that increased RMSSD, LF and HF (associated to decreased HR) were also observed post lockdown, is a marker of an effective and lasting improvement in cardiovascular fitness. The best performances occur when both the sympathetic and parasympathetic influences on the heart are high [12,13] hence increased LF and HF, associated to decreased resting HR.

There was a moderate but significant correlation between VAS and HRV, which was positive for HF and RMSSD and negative for HR. Therefore, the better the participants felt, the higher the parasympathetic influences and the lower the HR, and vice-versa the lower the HF and RMSSD, the lower the feeling of well-being. These later results emphasize the usefulness of HRV as a global index of heath in the general population. In line with previous literature and data from the national French public health institute, the more active people during the lockdown may have felt better [2,20].

### Limitations

The lockdown was applied suddenly, and it was therefore impossible to recruit participants a priori during the baseline period. By chance we had enough participants with sufficient HRV tests to conduct this study. Overall, it is a unique opportunity to describe the HRV behavior in 95 healthy people, extracted from a database of 7098 orthostatic tests. The main limitation is that we did not have any data on physical activity of these participants. We can only speculate that the HRV changes during CFN reflect at least partly the decrease or increase in physical activity.

## CONCLUSION

The lockdown in France was sudden and severe, drastically limiting the possibilities for physical activities. This likely resulted in a stressful situation and in a general cardiovascular detraining (i.e., decreased RMSSD and HF and increased resting heart rate in supine position) in most of the population. However, in 20% of the participants the lockdown was positively appreciated, as shown by an improved well-being and parasympathetic activity. Altogether, the present study confirms the usefulness of regular HRV testing in the general population for monitoring health status, as previously shown in elite athletes.

## Data Availability

Data available upon request to the authors

## Authors’ contributions

NB and GPM designed the study. NB and SY analyzed the data. SY did the signal processing. NB wrote the article and prepared the figures. GPM and LS reviewed the article. All the authors approved the final version of the manuscript and declare no conflict of interest.

## REFERENCES

1. Martinez-Ferran M, de la Guía-Galipienso F, Sanchis-Gomar F, Pareja-Galeano H. Metabolic Impacts of Confinement during the COVID-19 Pandemic Due to Modified Diet and Physical Activity Habits. Nutrients. 2020;12. doi:10.3390/nu12061549

2. Lemoine V, Delibéros M, Bessarion C, Champion S. Confinement?: un impact certain sur l’activité physique, le temps passé assis et le temps passé devant un écran. Press Release. 2020. doi:file:///C:/Users/n.bourdillon/Downloads/CP_CoviPrev_activite_physique_170620.pdf

3. Lavie CJ, Arena R, Swift DL, Johannsen NM, Sui X, Lee D-C, et al. Exercise and the cardiovascular system: clinical science and cardiovascular outcomes. Circ Res. 2015;117: 207–219. doi:10.1161/CIRCRESAHA.117.305205

4. Pareja-Galeano H, Sanchis-Gomar F, Lucia A. Physical activity and depression: type of exercise matters. JAMA Pediatr. 2015;169: 288–289. doi:10.1001/jamapediatrics.2014.3501

5. Schmitt L, Regnard J, Parmentier AL, Mauny F, Mourot L, Coulmy N, et al. Typology of “Fatigue” by Heart Rate Variability Analysis in Elite Nordic-skiers. Int J Sports Med. 2015;36: 999–1007. doi:10.1055/s-0035-1548885

6. Wang Y-P, Kuo TBJ, Li J-Y, Lai C-T, Yang CCH. The relationships between heart rate deceleration capacity and spectral indices of heart rate variability during different breathing frequencies. Eur J Appl Physiol. 2016;116: 1281–1287. doi:10.1007/s00421-016-3332-z

7. Holzman JB, Bridgett DJ. Heart Rate Variability Indices as Bio-Markers of Top-Down Self-Regulatory Mechanisms: A Meta-Analytic Review. Neurosci Biobehav Rev. 2017. doi:10.1016/j.neubiorev.2016.12.032

8. Akselrod S, Gordon D, Ubel FA, Shannon DC, Berger AC, Cohen RJ. Power spectrum analysis of heart rate fluctuation: a quantitative probe of beat-to-beat cardiovascular control. Science. 1981;213: 220–222.

9. Goldstein DS, Bentho O, Park M-Y, Sharabi Y. Low-frequency power of heart rate variability is not a measure of cardiac sympathetic tone but may be a measure of modulation of cardiac autonomic outflows by baroreflexes. Exp Physiol. 2011;96: 1255– 1261. doi:10.1113/expphysiol.2010.056259

10. Pomeranz B, Macaulay RJ, Caudill MA, Kutz I, Adam D, Gordon D, et al. Assessment of autonomic function in humans by heart rate spectral analysis. Am J Physiol. 1985;248: H151–153.

11. Dixon EM, Kamath MV, McCartney N, Fallen EL. Neural regulation of heart rate variability in endurance athletes and sedentary controls. Cardiovasc Res. 1992;26: 713– 719.

12. Hug B, Heyer L, Naef N, Buchheit M, Wehrlin JP, Millet GP. Tapering for marathon and cardiac autonomic function. Int J Sports Med. 2014;35: 676–683. doi:10.1055/s-0033-1361184

13. Parouty J, Al Haddad H, Quod M, Leprêtre PM, Ahmaidi S, Buchheit M. Effect of cold water immersion on 100-m sprint performance in well-trained swimmers. Eur J Appl Physiol. 2010;109: 483–490. doi:10.1007/s00421-010-1381-2

14. Buchheit M. Monitoring training status with HR measures: do all roads lead to Rome? Front Physiol. 2014;5: 73. doi:10.3389/fphys.2014.00073

15. Teisala T, Mutikainen S, Tolvanen A, Rottensteiner M, Leskinen T, Kaprio J, et al. Associations of physical activity, fitness, and body composition with heart rate variability-based indicators of stress and recovery on workdays: a cross-sectional study. J Occup Med Toxicol Lond Engl. 2014;9: 16. doi:10.1186/1745-6673-9-16

16. Bourdillon N, Schmitt L, Yazdani S, Vesin J, Millet GP. Minimal Window Duration for Accurate HRV Recording in Athletes. Front Neurosci. 2017;11: 456.

17. Thorpe RT, Strudwick AJ, Buchheit M, Atkinson G, Drust B, Gregson W. Tracking Morning Fatigue Status Across In-Season Training Weeks in Elite Soccer Players. Int J Sports Physiol Perform. 2016;11: 947–952. doi:10.1123/ijspp.2015-0490

18. Lavie CJ, Ozemek C, Carbone S, Katzmarzyk PT, Blair SN. Sedentary Behavior, Exercise, and Cardiovascular Health. Circ Res. 2019;124: 799–815. doi:10.1161/CIRCRESAHA.118.312669

19. Ozemek C, Laddu DR, Lavie CJ, Claeys H, Kaminsky LA, Ross R, et al. An Update on the Role of Cardiorespiratory Fitness, Structured Exercise and Lifestyle Physical Activity in Preventing Cardiovascular Disease and Health Risk. Prog Cardiovasc Dis. 2018;61: 484–490. doi:10.1016/j.pcad.2018.11.005

20. Brooks SK, Webster RK, Smith LE, Woodland L, Wessely S, Greenberg N, et al. The psychological impact of quarantine and how to reduce it: rapid review of the evidence. Lancet Lond Engl. 2020;395: 912–920. doi:10.1016/S0140-6736(20)30460-8

21. Matias T, Dominski FH, Marks DF. Human needs in COVID-19 isolation. J Health Psychol. 2020;25: 871–882. doi:10.1177/1359105320925149

22. Jiménez-Pavón D, Carbonell-Baeza A, Lavie CJ. Physical exercise as therapy to fight against the mental and physical consequences of COVID-19 quarantine: Special focus in older people. Prog Cardiovasc Dis. 2020. doi:10.1016/j.pcad.2020.03.009

23. Burtscher J, Burtscher M, Millet GP. (Indoor) isolation, stress and physical inactivity: vicious circles accelerated by Covid-19? Scand J Med Sci Sports. 2020. doi:10.1111/sms.13706

